# Outcomes of Endocarditis in Pregnancy: A Single Center Experience

**DOI:** 10.1101/2023.05.05.23289602

**Authors:** Kayle Shapero, Kathryn Berlacher, Christina Megli

## Abstract

**Background:** The incidence of infective endocarditis (IE) in pregnancy is rare (0.006%) with increasing prevalence during the opioid epidemic. IE in pregnancy is associated with high rates of maternal and fetal mortality and morbidity, and existing data on outcomes in pregnancy is limited to case reports and small studies. Our study compares the outcomes of pregnant patients with IE as compared to non-pregnant patients.

**Methods:** Patients diagnosed with IE during pregnancy or 30 days post-partum between 2014-2021 were identified by ICD-9/10 codes. Pregnant cases were matched to a random sampling of non-pregnant reproductive-age female endocarditis patients in a 1:4 ratio. Demographic and clinical data was collected. Data was validated through chart review.

**Results:** 180 patients with IE were identified for this cohort; 34 were pregnant or within 30 days post-partum at diagnosis. There were higher rates of hepatitis C and opioid maintenance therapy in the pregnant patients. Etiology of IE in pregnant patients was predominantly *S. aureus* (MSSA/MRSA) whereas non-pregnant woman had greater microbiological variation. We observed comparable rates of valve replacement (32.4% vs 29% p=0.84), 2-yr mortality (20.6% vs 17.8%, p =.>0.99), and ICU admission (61.8% vs 69.8%, p=0.414) in pregnant patients. There was a trend towards higher rates of PE (17.6% vs 7.5%, p=0.098), arrhythmia (17.6% vs 9.6%, p=0.222) among pregnant patients. There were high rates of IVDU relapse in both groups (>40%).

**Conclusion:** We observed similar rates of valve replacement, 2-year mortality, and ICU admission in both pregnant and non-pregnant patients with IE. We observed a microbial predilection for *S*.*aureus* in pregnancy suggesting that the physiology of pregnancy may select for this microbiologic etiology. These results represent the largest single center retrospective review of IE in pregnancy and suggests that pregnancy may have increased morbidity.

## Introduction

Cardiovascular disease and infection are the leading causes of maternal mortality and severe morbidity, with infective endocarditis (IE) in pregnancy representing the intersection of these diseases. While the incidence of IE in pregnancy has historically been reported as rare (0.006%)^1^, increasing incidence in pregnant and non-pregnant patients have been reported paralleling the ongoing opioid epidemic c.^2,3^ Infectious morbidity due to injection drug use is an under-evaluated consequence of the opioid epidemic as persons who inject drugs are at increased risk for severe bacterial infections leading to high costs and high morbidity. The majority of cases of IE in pregnancy are now attributable to opioid use disorder.^2,3^ The physiologic and immune adaptations to pregnancy may increase the risk for complications and adverse outcomes. Additionally, the consideration of both maternal and fetal well-being can add additional complexities to care.^4^ Existing data on outcomes of IE in pregnancy is primarily limited to case reports and case series, and there are no clear recommendations regarding optimal management.

The cardiovascular adaptations to pregnancy confer a substantial risk of hemodynamic compromise due to valvular disease. Exact mortality data for IE in pregnancy is unknown, with reports ranging from 5-33%.^1,2^ Recent studies demonstrate improving outcomes, likely due to early recognition and treatment^1,3^. Treatment ranges from medical management to surgical intervention depending on the severity of disease and infectious complications, although in pregnancy the ideal treatment is unknown. For those patients with significant hemodynamic compromise prompting consideration of extracorporeal membrane oxygenation, there has been recent limited evidence that it does not confer excess maternal risk in pregnancy.^5^ While maternal outcomes from cardiac surgery are *reportedly* similar to non-pregnant female patients, current literature is limited to case series and retrospective reviews. The use of cardiopulmonary bypass and cardiac surgery in pregnancy is still controversial, and data on rates of surgical intervention are limited.^2^ Management decision regarding timing of valve surgery are complex given poor fetal outcomes reported with cardiopulmonary bypass^6^.

Given the lack of current data, we performed a retrospective single center cohort study of IE outcomes in pregnant patients as compared to their non-pregnant counterparts to assess the impact of pregnancy on IE outcomes. Our study defines the outcomes of pregnant patients with IE as compared to non-pregnant patients and furthermore identifies antenatal characteristics associated with maternal and neonatal morbidity and mortality.

## Methods

### Patient Population

We limited our study to hospital admissions that occurred between 2014 and 2021 at the primary obstetric hospital in a large health care system. These dates were selected based on data availability and ensured the use of both the ninth and tenth revision of the International Classification of Diseases, Clinical Modifications (ICD), and the 10^th^ edition adopted in October 2015. Patients were included in the study if they were female sex, reproductive age (18-45), had at least one ICD code indicating IE and had an encounter code for an echocardiogram within 6 months of IE diagnosis (see Supplement). We excluded patients with IE diagnosis and treatment occurring at satellite facilities. For patients with multiple IE admissions, the first index admission within the system during that time frame was used. We included those patients who were treated for IE according to Duke’s criteria ^7^ even if no echocardiographic evidence of endocarditis was identified. Within this cohort of patients, chart review was performed to identify those who were pregnant or within 30 days postpartum at the time of IE diagnosis. Patient data was validated through chart review of a random sampling of non-pregnant female patients with IE of reproductive age (between the ages of 18-45) in a 1:4 ratio.

### Definitions

For comparison of the effect of pregnancy on IE, the following the outcomes were included: mortality (in-hospital, 1 and 2-year), valve replacement, valve repair, other valve surgeries, thromboembolic event (including stroke, pulmonary embolism, septic pulmonary emboli), arrhythmia, intubation, use of mechanical support, use of ECMO, ICU admission, length of ICU and hospital stay. Of note, leukocytosis/leukopenia in pregnancy was defined by the reference values during each trimester of pregnancy (first trimester 5.7x10^9^/L to 13.6x10^9^/L, second trimester 5.6x10^9^/L to 14.8x10^9^/L, third trimester 5.9x10^9^/L to 16.9x10^9^/L).^8^ A complete list of outcomes can be found in Supplementary Table 2. Neonatal outcomes were calculated using the denominator of patients that remained pregnant until at least 23weeks gestation age with a viable intrauterine pregnancy at that time.

### Statistical analysis

Demographic, clinical and maternal outcomes related variables were compared. Statistical analysis was done using STATA. Categorical variables were analyzed using fisher’s exact t-tests. Continuous variables were compared using t-test or Mann-Whitney test (if data did not follow a normal distribution). This study was approved via the University of Pittsburgh Medical Center Internal Review Board (STUDY21060177).

## Results

### Population Characteristics

During our study period, we identified 34 pregnant or recently postpartum patients and matched them to 146 controls with a diagnosis of IE and an incident echocardiogram. The majority of pregnancy-associated IE occurred during index admission for delivery (76.5%) and the remainder occurred post-partum.

Pregnant patients with IE were significantly younger than non-pregnant patients (29.6 vs 32.6; P=0.011). There was no significant difference between percentage of patients on Medicaid or distance to tertiary hospital, although there were more Caucasian patients in the pregnant cohort (97% vs 85.6%, p=0.08). There were higher rates of hepatitis C (68% vs. 42%, p=0.012) in pregnant patients. There were lower rates of comorbidities of hypertension, hyperlipidemia, coronary artery disease, chronic kidney disease, diabetes and asthma among pregnant patients, although these differences did not reach statistical significance. Rates of mental health disorders were high in both groups, 36% in non-pregnant and 44% in pregnant patients, with depression, anxiety and bipolar disorder as the most prevalent disorders. There was a trend towards significance in the higher rates of incarceration noted in pregnant as compared to non-pregnant patients. In terms of risk factors, patients with pregnancy associated IE had higher rates of native valve predisposition including congenital heart disease (11.8%), bicuspid aortic valve (5.8%) and mitral valve prolapse (2.9%). However, a greater number of patients with non-pregnancy associated IE had a history of IE (27.4% vs 17.7%; p= 0.28) and presence of prosthetic valve (10=1% vs 2.9%; p =0.343). Intravenous drug use (85.3% vs 80.8%, p=0.63) and opioid maintenance therapy (58.5% vs 24%, p=0.000) was higher among pregnant patients.

### Incident presentation

The majority of symptoms on presentation were similar between both groups including nonspecific symptoms of fevers, chills, myalgias, back pain, and chest pain. However, we noted increased incidence of shortness of breath among pregnant patients (41.% vs 11%, p=0.000), and decreased incidence of stroke/altered mental status symptoms (5.9% vs 21.3% p = 0.047) as compared to non-pregnancy related IE. Of note, 17.6% (n=6) of the pregnant patients ultimately diagnosed with IE presented for non-infectious reasons (ie in labor, methadone conversion, trauma), similar to the number of non-pregnant patients who presented for non-infectious reasons (22.6%). Symptom duration prior to presentation, and presence of fevers was similar between both groups, although the incidence of leukocytosis was higher in the non-pregnant population (61% vs 35%, p=0.007). Both groups had rapid attainment of blood cultures (<24hrs) and similar rates of screening for hepatitis C (50-60%). There were higher rates of urine drug screens among pregnancy associated IE (82.4% vs 50.7%, p=0.001).

### Endocarditis details

Approximately 50% of IE cases affected the tricuspid valve, although the incidence of right and left sided lesions was split evenly in both groups. Incidence of bioprosthetic valve endocarditis was higher in non-pregnant patients, due to a greater incidence of prosthetic valves, but did not reach significance. Fewer transesophageal echocardiograms were performed on pregnant patients as compared to non-pregnant patients (50% vs 69.8% p=0.043) likely due to concerns regarding fetal exposure to anesthesia.

### Microbial Variation

Etiology of IE in pregnant women was predominantly *S. aureus* (MSSA/MRSA) whereas in non-pregnant woman IE had greater microbiological variation (82% vs. 65%, p= 0.03) (Figure 1).

**Figure 1a.**
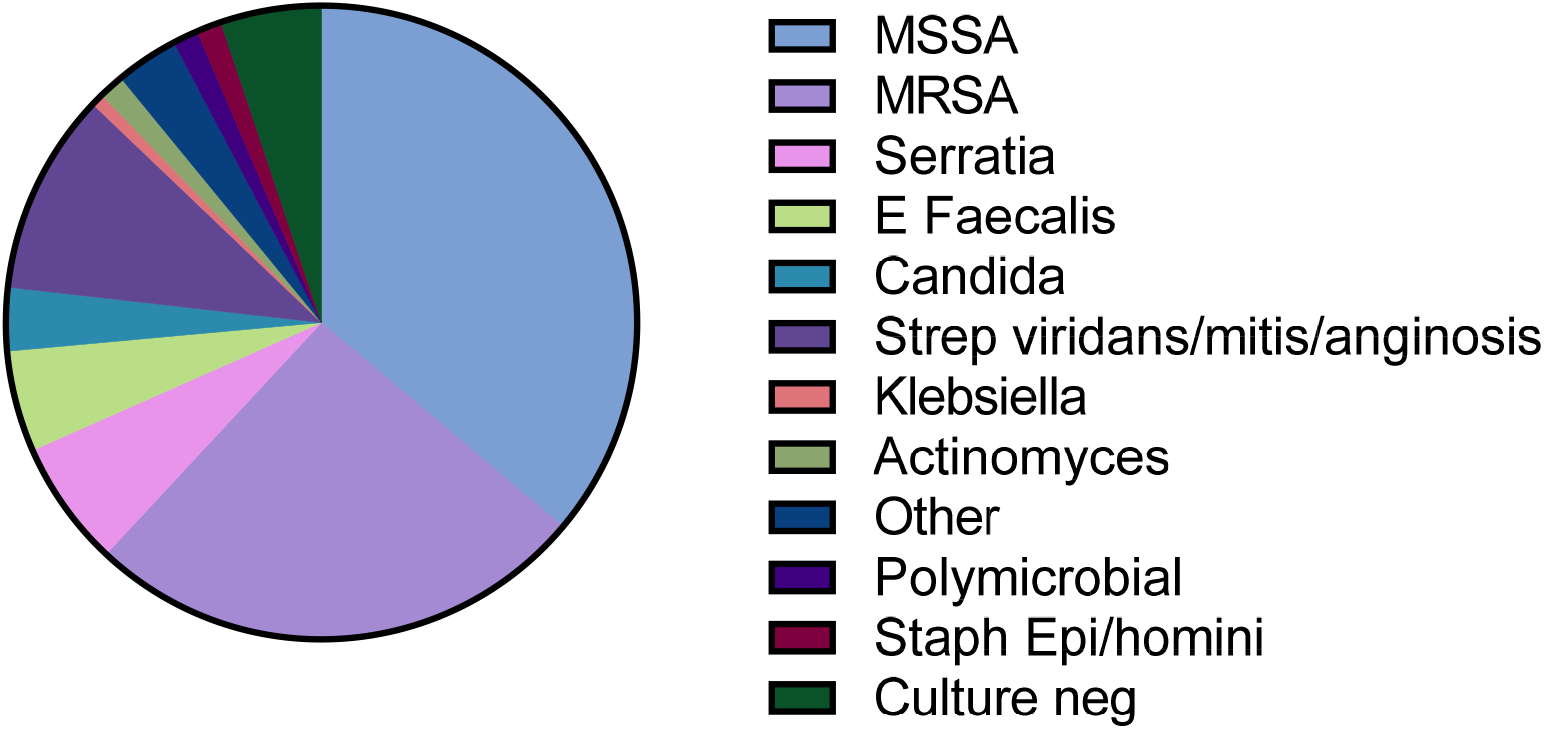
Infectious Agent- Non-Pregnant patients.

**Figure 1a.**
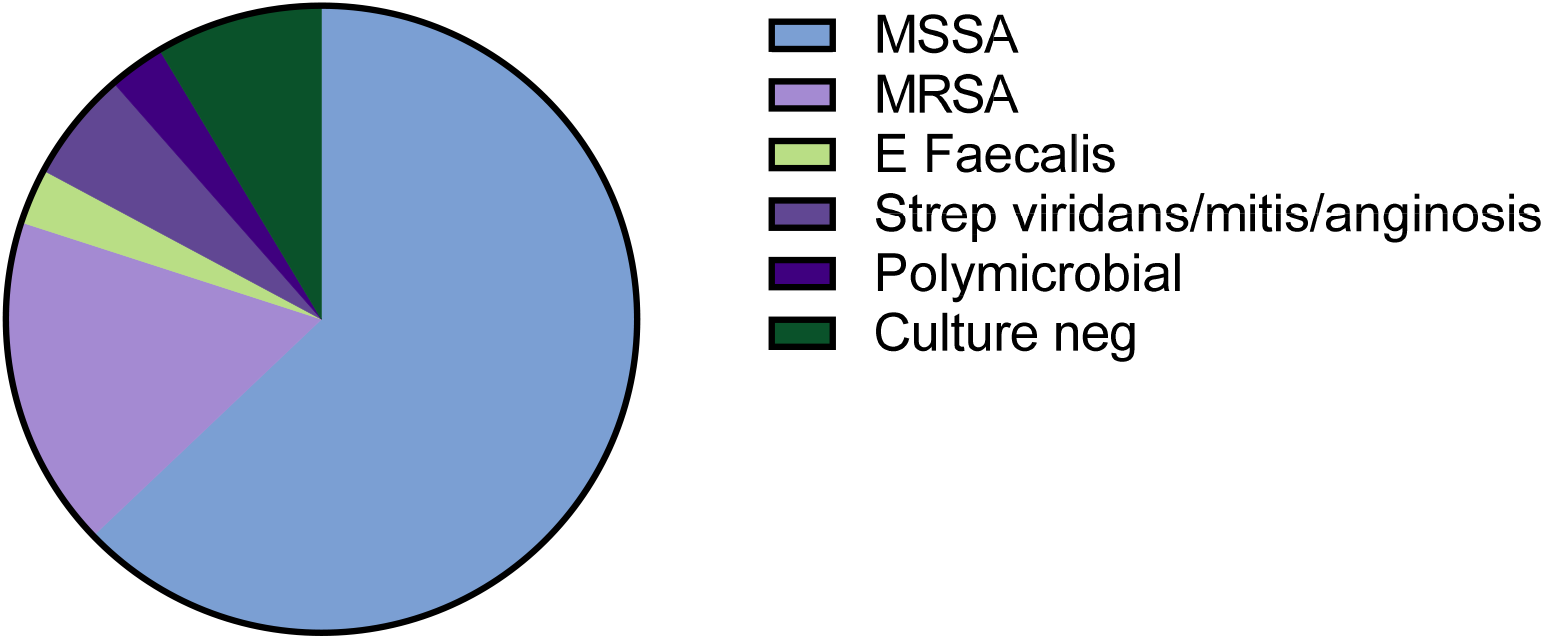
Infectious Agent- Pregnant patients.

### Maternal Outcomes

Maternal outcomes of pregnancy-associated and non-pregnancy associated IE are detailed in Table 3. We found 1 year (15.1% vs 14.7%) and 2-year (17.8% vs 20.6%) mortality to be similar between both groups. There was increased mortality early in the disease course in pregnant patients (in-hospital mortality9.6% vs 2.9%). We observed similar rates of patients having 1 or more surgical indications for valvular surgery (68.5% vs 73.5%, p=0.681), (as set forth by 2014 ACC/AHA guidelines on the Management of Patients with Valvular Heart Disease^9^). Despite 73.5% of patients with indications, no surgeries were performed during pregnancy. Total rates of valve replacement (32.4% vs 29.5% p=0.836) and valve repair were similar between groups (5.5% vs 5.9%, p>0.999). When adjusted for the total number of patients meeting surgical indications, rates of intervention remained similar (51% vs 52%, p=>0.999). With regards to IE complications, we observed a trend towards higher rates of other septic emboli (including splenic/hepatic infarcts, renal infarcts, septic arthritis, empyema and abscesses) in non-pregnancy related IE (52.1% vs 35.3% p = 0.089). Despite a trend toward increased rates of septic emboli in non-pregnant IE, pregnancy was associated with increased persistence despite appropriate antibiotic treatment. We also observed higher rates of pulmonary embolus in pregnancy related IE, although this did not reach statistical significance (17.6% vs 7.5%, p=0.098). We observed similar rates of renal failure, cardiac arrest, ARDS, heart failure, arrhythmia and ICU admission among both groups. For those who underwent surgical replacement or repair, total cardio-pulmonary bypass time and the time duration from admission to surgical intervention did not differ between groups. Additionally, rates of post-surgical complications including the need for temporary pacing wires, defibrillator placement, and intracranial-hemorrhage were similar between both groups. There were high rates of 30-day readmission (>20%) and IVDU relapse in both groups (51.7% in non-pregnant and 45.2% in the pregnant patients).

**Table 1.** Demographics.

|  | Non-Pregnant Average |  | Pregnant Average |  | P-Value |
| --- | --- | --- | --- | --- | --- |
|  | Median or Count | Percentage or IQR/SD | Median or count | Percentage or IQR/SD |  |
| **<u>Demographics</u>** |  |  |  |  |  |
| **Age at delivery** | 32.6 | 6.9 | 29.6 | 5.8 | 0.011 |
| **Race** |  |  |  |  |  |
| White | 125 | 85.6 | 33 | 97.1 | 0.082 |
| Black | 12 | 8.2 | 1 | 2.9 | 0.467 |
| American Indian | 1 | 0.7 | 0 | 0 | >0.99 |
| Unspecified/declined | 10 | 6.9 | 0 | 0 | 0.212 |
| **Medicaid/medicare** | 105 | 71 | 26 | 76.5 | 0.673 |
| **<u>Comorbidities</u>** |  |  |  |  |  |
| **HTN** | 23 | 15.8 | 3 | 8.8 | 0.42 |
| **HLD** | 6 | 4.1 | 0 | 0 | 0.596 |
| **CAD** | 1 | 0.7 | 0 | 0 | >0.99 |
| **CKD** | 10 | 6.6 | 0 | 0 | 0.212 |
| **Obesity** | 8 | 5.5 | 1 | 2.9 | >0.99 |
| **CVA/dvt** | 14 | 9.6 | 3 | 8.8 | >0.99 |
| **Hepatitis C** | 62 | 42.5 | 23 | 67.6 | 0.012 |
| **DM** | 17 | 11.6 | 1 | 2.9 | 0.203 |
| **Thyroid disease** | 3 | 2.1 | 1 | 2.9 | 0.571 |
| **Asthma** | 16 | 11.0 | 3 | 8.8 | >0.99 |
| **Smoking history** | 102 | 69.9 | 26 | 76.4 | 0.532 |
| **<u>IVDU</u>** | 118 | 80.8 | 31 | 91.2 | 0.208 |
| **<u>Specific substance</u>** |  |  |  |  |  |
| **Mental health d/o** | 53 | 36.3 | 15 | 44.1 | 0.435 |
| **Incarceration** | 3 | 5.7 | 3 | 8.8 | 0.082 |
| **<u>Endocarditis risk factors</u>** |  |  |  |  |  |
| **<u>Native valve predisposition</u>** |  |  |  |  |  |
| Degen heart disease | 1 | 0.7 | 0 | 0 | >0.99 |
| Bicuspid Aortic Valve | 6 | 4.1 | 2 | 5.8 | 0.596 |
| Mitral valve prolapse | 1 | 0.7 | 1 | 2.9 | 0.343 |
| **Prosthetic valve** | 19 | 13.0 | 2 | 5.9 | 0.375 |
| **Intracardiac devices (ICD/Defib)** | 2 | 1.4 | 0 | 0 | >0.99 |
| **Congenital heart disease** | 11 | 7.5 | 4 | 11.8 | 0.49 |
| **History of endocarditis** | 40 | 27.4 | 6 | 17.7 | 0.28 |
| **IVDU** | 118 | 80.8 | 29 | 85.3 | 0.63 |
| **Chronic IV access** | 6 | 4.1 | 1 (2.9) | 2.9 | >0.99 |
| **<u>Protective risk factors:</u>** |  |  |  |  |  |
| **Opiate maintenance therapy** | 35 | 24.0 | 20 | 58.8 | >0.001 |
| Methadone | 11 | 7.5 | 7 | 20.6 | 0.05 |
| Suboxone | 24 | 16.4 | 13 | 38.2 | 0.008 |

**Table 2.** Infectious Details.

|  | Non Pregnant Average |  |  | Pregnant |  |  | P-value |
| --- | --- | --- | --- | --- | --- | --- | --- |
|  | Median or Count | Percentage | IQR/ SD | Median or count | Percentage | IQR/ SD |  |
| **Fevers** | 48 | 32.9 |  | 16 | 47.1 |  | 0.163 |
| **Chills** | 20 | 13.7 |  | 5 | 14.7 |  | >0.999 |
| **SOB** | 17 | 11. |  | 14 | 41.2 |  | <0.001 |
| **Myalgias** | 21 | 14.4 |  | 2 | 5.9 |  | 0.257 |
| **Back pain** | 14 | 9.6 |  | 2 | 5.9 |  | 0.740 |
| **CP** | 11 | 7.5 |  | 3 | 8.8 |  | 0.730 |
| **CVA symptoms/AMS** | 31 | 21.3 |  | 2 | 5.9 |  | 0.047 |
| **Other** | 33 | 22.6 |  | 7 | 20.6 |  | >0.999 |
| **Symptom duration prior to presentation** | 5 |  | (2,10) | 6 |  | (2.2,7.75) | 0.170 |
| **Febrile on admission** | 66 | 45.2 |  | 13 | 38.2 |  | 0.566 |
| **Leukocytosis/leukopenia on admission** | 89 | 61.0 |  | 12 | 35.2 |  | 0.007 |
| **Time to first blood cultures** |  |  |  |  |  |  |  |
| **<24hr** | 140 | 95.9 |  | 32 | 94.2 |  | 0.647 |
| **>24hr** | 6 | 4.1 |  | 2 | 5.9 |  | 0.647 |
| **Screened for hep C** | 78 | 53.4 |  | 22 | 64.7 |  | 0.256 |
| **AB** | 60 | 41.1 |  | 18 | 52.9 |  | 0.250 |
| **PCR** | 23 | 15.8 |  | 9 | 26.5 |  | 0.144 |
| **Screened if not known Hepatitis C** | 40 | 47.6 |  | 8 | 72.7 |  | 0.017 |
| **TEE performed (yes/no)** | 102 | 69.8 |  | 17 | 50.0 |  | 0.043 |
| **Time from admission to TEE** | 3 |  | (2,4) | 3 |  | (1,10) | 0.529 |
| **UDS** | 74 | 50.7 |  | 28 | 82.4 |  | 0.001 |
| **Left AMA** | 25 | 17.1 |  | 8 | 23.5 |  | 0.460 |
| **<u>Endocarditis details:</u>** |  |  |  |  |  |  |  |
| **Valve location** |  |  |  |  |  |  |  |
| **Aortic** | 26 | 17.8 |  | 5 | 14.7 |  | 0.804 |
| **Mitral** | 42 | 28.8 |  | 12 | 35.3 |  | 0.534 |
| **Pulmonic** | 3 | 2.1 |  | 2 | 5.9 |  | 0.239 |
| **Tricuspid** | 76 | 52.1 |  | 16 | 47.1 |  | 0.704 |
| **No valve** | 14 | 9.6 |  | 5 | 14.7 |  | 0.364 |
| **Type of valve** |  |  |  |  |  |  |  |
| **Native** | 128 | 87.7 |  | 28 | 82.4 |  | 0.408 |
| **Prosthetic** | 19 | 13.0 |  | 2 | 5.9 |  | 0.375 |
| **Duration of bacteremia (days)** | 5.1 |  | (2,7) | 5.9 |  | (3,7.5) | 0.261 |
| **Valvular details** |  |  |  |  |  |  |  |
| **Regurgitation severity** |  |  |  |  |  |  |  |
| **Mild regurgitation** | 37 | 25.3 |  | 9 | 26.5 |  | 0.681 |
| **Mod regurgitation** | 33 | 22.6 |  | 9 | 26.5 |  | 0.655 |
| **Severe regurgitation** | 56 | 38.4 |  | 10 | 29.4 |  | 0.430 |
| **No regurgitation** | 20 | 13.7 |  | 6 | 17.6 |  | 0.590 |
| **EF on TTE/TEE (%)** | 57.6 |  | (55,60) | 56.3 |  | (55,57.5) | 0.313 |
| **Vegetation size** |  |  |  |  |  |  |  |
| **1-dimension** | 1.8 |  | (1.1,2.4) | 1.9 |  | (1.0,2.4) | 0.863 |
| **2-dimension** | 1.0 |  | (0.58,1.4) | 1.0 |  | (0.63,1.55) | 0.329 |
| **>1 vegetation** | 4 | 2.7 |  | 5 | 14.7 |  | 0.013 |
| **No veg** | 21 | 14.4 |  | 6 | 17.6 |  | 0.601 |
| **Suspected source** |  |  |  |  |  |  |  |
| **IVDU** | 113 | 77.4 |  | 25 | 73.5 |  | 0.655 |
| **Other** | 22 | 15.1 |  | 9 | 26.5 |  | 0.131 |
| **Duration of antimicrobial treatment (weeks)** | 6.1 |  | (6,6) | 5.8 |  | (6.6) | .6910 |
| **Successful completion of antibiotic therapy** | 69 | 47.3 |  | 20 | 58.8 |  | 0.256 |
| **Antiplatelet therapy** | 5 | 3.4 |  | 0 | 0.0 |  | 0.585 |

**Table 3:**
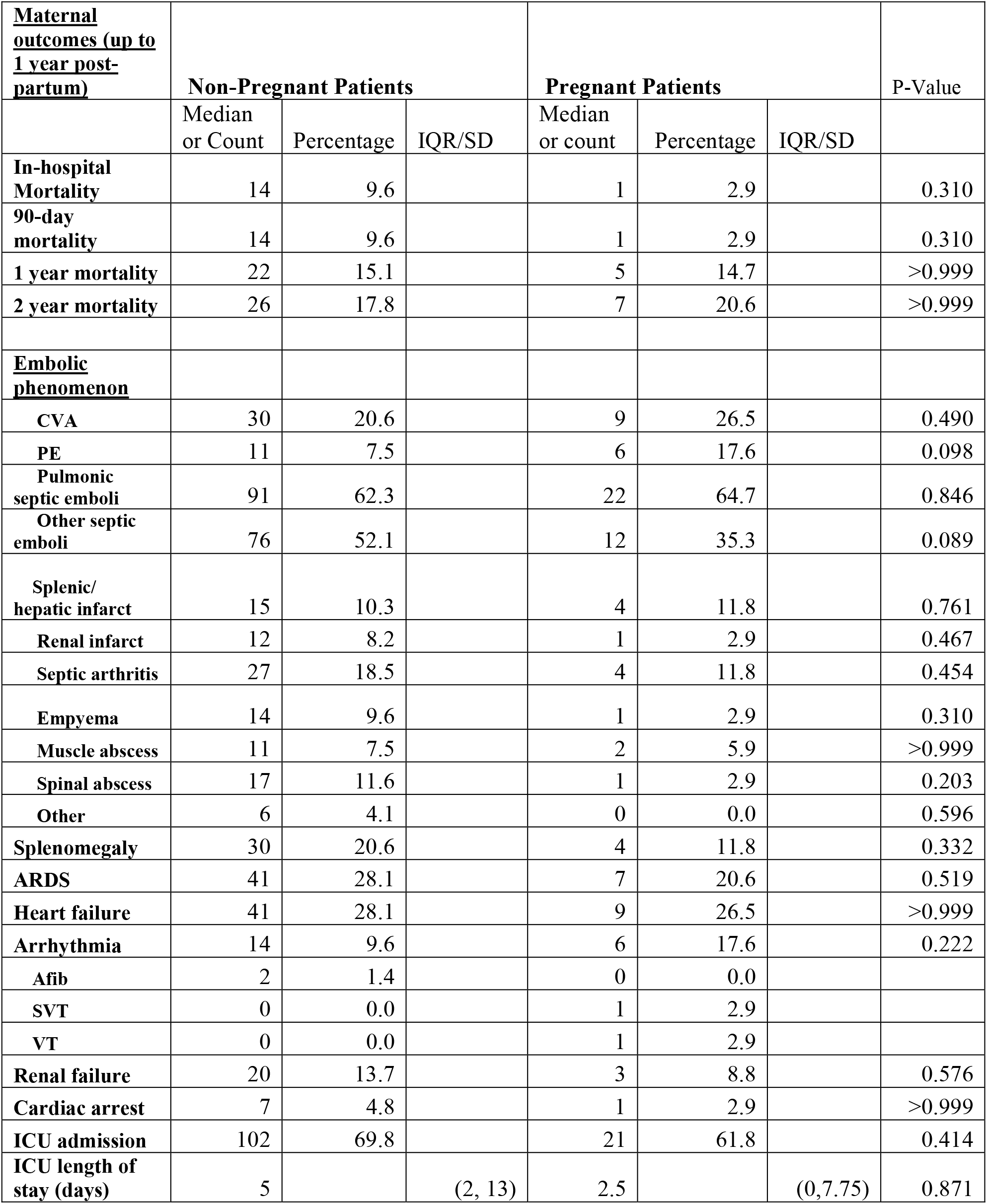
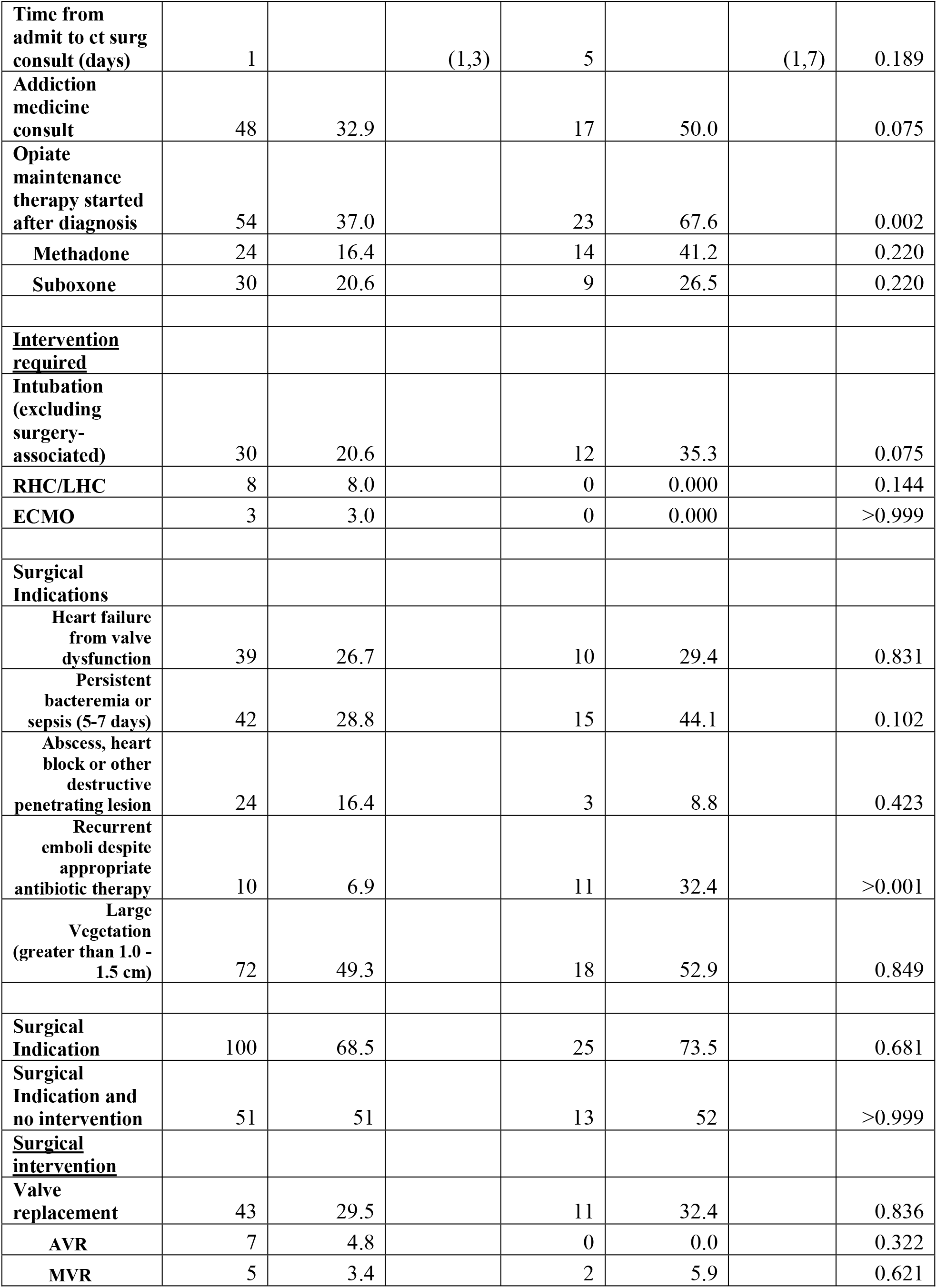
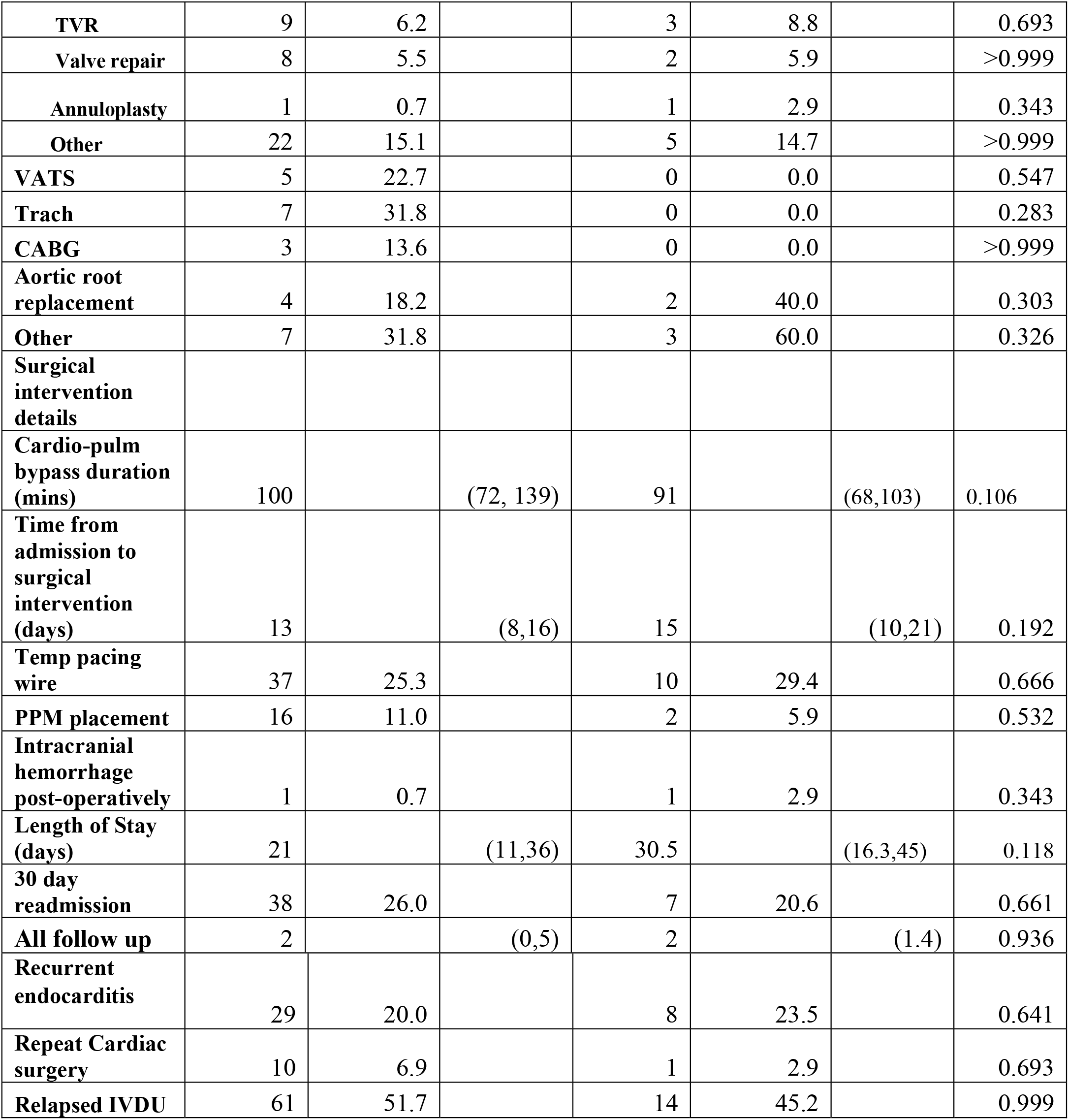
Maternal Outcomes.

### Delivery and Neonatal outcomes

The delivery and neonatal outcomes are detailed in Table 4. Of available delivery outcomes, there were 26 live births (76.4%), 3 (9%) terminations and 4 (15.4%) spontaneous abortions. Of patients that were expectantly managed, perinatal mortality (including intrauterine fetal demise and previable delivery) was high at 15.4%. There was a very high preterm delivery rate of 50% (n = 11) with average live gestational age at birth of 35.0 weeks. Accordingly, the majority of infants 68% required NICU admission.

**Table 4.** Delivery and Fetal Outcomes.

| Delivery and Fetal outcomes | Total N=34 | Percentage |
| --- | --- | --- |
| Intrapartum IE | 29 | 85.3% |
| Post Partum IE | 5 | 14.7% |
| Live births | 26 | 70% |
| Perinatal mortality | 4 | 15.4% |
| Termination | 3 | 9% |
| Mode of Delivery |  |  |
| C-section | 9 | 41% |
| Spontaneous vaginal delivery | 19 | 86% |
| c-section indication for maternal instability | 3 | 14% |
| Induction of labor | 6 | 27.3% |
| Average gestational age of live birth | 35w |  |
| Preterm delivery | 11 | 50% |
| NICU stay | 15 | 68% |

## Discussion

The incidence rate of IE, particularly among patients with opioid use disorder has increased nearly ten-fold in the last decade, from 3.7 in 2011 to 30.1 per day per one million people in 2022.^10^ This epidemic also affects women of child bearing age, and as such the rates of IE in pregnancy are estimated to be 0.006%.^1^ There has been conflicting data regarding overall mortality and morbidity in this population. Our study evaluated a cohort of pregnant and post-partum patients with IE and compared outcomes to a non-pregnant cohort and found mortality to be high and major complication rates to be low and equivalent in both groups. We observed similar 1-year mortality rates (15%) in both groups, as well as similar rates of complications such as arrhythmias, cardiac arrest, acute respiratory distress syndrome and renal failure. This result is in line with a recent analysis of the National Readmissions Database which found that pregnancy did not confer an increased risk of IE associated mortality and was not associated with increased adverse outcomes. These combined results suggest that we need to reconsider our approach to treating these patients. While pregnant patients with IE have been considered extremely high risk, with many providers reticent to intervene surgically, our study suggests that they would benefit from the same aggressive intervention as non-pregnant IE patients.

With regards to maternal outcomes, we did not observe any significant differences in rates of valve repair or replacement between the two groups. It is established that the use of cardio-pulmonary bypass confers higher fetal risk and the incidence of cardiac surgery in pregnancy is quite limited^2,11^. As such, although we observed similar rates of surgical intervention (30%) in both groups. However, it is important to clarify that all surgeries in the pregnancy associated IE patients were performed after delivery and patients with an antenatal diagnosis of IE were admitted until delivery (admission lengths 16-45days) and for treatment and coordination of care postpartum. This suggests that although surgery was avoided during pregnancy, the added hemodynamic challenges of the post-partum state did not significantly impact the decision to operate. The absence of any statistically significant difference in outcomes or surgical complications suggests that cardiac surgery can be effectively and safely pursued in the post-partum period, and should not be deferred by weeks or months. Additionally, the introduction of a multidisciplinary endocarditis team at our institution in 2017 may have improved rates of intervention among pregnant patients.

While rates of mortality in particular were similar between pregnant and non-pregnant patients, the one-year mortality rate of 15% is still unacceptably high in this young and otherwise healthy population. A secondary goal of this study was to identify factors related to these poor outcomes and identify opportunities for intervention. The observed increase in IE rates has been driven in part by the rise in intravenous drug use, including both opiates and stimulants.^12^ Unsurprisingly, in this study we observed high rates (>80%) of IVDU in both groups. Despite the known concurrence of IVDU and IE, in this study we observed suboptimal rates of urinary drug screening in both pregnant and non-pregnant patients. The American College of Obstetrics and Gynecology (ACOG) recommends using a questionnaire to screen all pregnant women regarding drug use, however it does not recommend routine urine drug screens. Despite these recommendations there is existing data that pregnant women presenting to the ED are less likely to be tested for ETOH or drug use than non-pregnant patients^13^ which is in direct contrast to our results. Similarly, with regards to Hep C, disappointingly only 47.6% of the non-pregnant and 72.7% of the pregnant patients without known hep C were screened with either Ab or PCR on presentation. This is surprisingly low, despite the known very high prevalence of anti HVC antibodies among IV drug users in the US (approximately 70%). ^14^

Despite increased rates of opioid use disorder screening and availability of treatment, data suggests that large percentages of hospitalized pregnant patients do not receive medication for opioid use disorder. Historically patients with injection drug use and/or pregnancy have high rates of patient-directed discharges and failure of antibiotic completion.^15^ Screening and treatment implementation for opioid use disorder in pregnant patients with IE represents another area lacking in robust data. With the rising rates of OUD and correlation with infectious endocarditis incidence, treatment of OUD offers another opportunity for multidisciplinary teams to directly impact outcomes in the pregnant population.^4^ In our population although screening for substance use was suboptimal in the non-pregnant population (as discussed above), in general we did observe higher rates of consultation for addiction medicine and higher rates of discharge on opiate replacement therapy (statistically significant in pregnant patients). Given the data that use of additional medicine consultation can lower readmission rates^16^ this offers an opportunity for interventions such as increased OUD screening and increased implementation of OUD treatment. In addition to OUD, high rates of concurrent mental health disorders have also been observed in patients with IV drug use, both in this study as well as in the literature.^17^ Interestingly, in our study we noted a trend towards increased rates of incarceration in pregnant patients, which is likely due to a Pennsylvania policy requiring methadone conversion for incarcerated pregnant patient with OUD.^18^ Ensuring timely, accurate, and consistent diagnosis of mental health disorders as well as providing options for treatment represent additional opportunities for intervention.

Our study has highlight several possible avenues of intervention which could impact diagnosis and treatment of this vulnerable patient population, including enhanced OUD and mental health disorder screening, and implementation of OUD treatment. In addition, the identification of staph aureus as the primary etiology of IE in the pregnant patients suggests a that pregnancy may somehow select for this subset of infection. Additionally, the knowledge that the majority of infections are due to staph aureus could raise the index of suspicion and increase the utility of a thorough and detailed exam to help with early identification of patients with IE.

While our study focused primarily on maternal outcomes in IE it is important to note that data on fetal outcomes is also sparse in the IE literature. One of the most commonly cited complications is high preterm delivery rate, with the average gestational age at live birth of 32 weeks.^19^ This can lead to further neonatal complications requiring ICU admissions, prolonged lengths of stay, as well as increase in the incidence of neonatal abstinence syndrome. These fetal complications have been associated with adverse neurodevelopmental outcomes and higher disability rates in children, however the long-term pediatric outcomes related to maternal endocarditis have not been studied.^20^ Similar to what has been previously described, we observed high rates of fetal mortality (21.5% and high rates of preterm delivery (46%), which is in line with most recent fetal outcomes^21^

There are many strengths of this study. As mentioned, the previous literature describing maternal outcomes in IE have largely been limited to case reports with the exception of a recent analysis of the National Readmissions Database assessing adverse outcomes of IE in pregnancy. It is notable that the outcomes examined in the prior study were mortality, thromboembolic events, and ventilation, and were largely limited by the database itself. Given that our study involved primary chart review, we were able to include far more detailed analysis of hospital course, complications and presentation. Additionally, this study represents one of the largest single center analyses of IE in pregnancy. There are also several limitations of this study worth noting. Given the relative rarity incidence of IE in pregnancy, the total sample size of pregnancy associated IE is quite small at 34 patients, which limits the power of our statistical analysis.

This study represents the largest known single center retrospective review of endocarditis in pregnancy and suggest that pregnancy does not portend an increased risk of overall mortality and morbidity in this population of patients. Although the incidence of IE in pregnancy remains rare, it is an increasingly common complication as the rates of opioid use disorder rapidly rise in this country. This study enriches the sparse literature surrounding such a vulnerable population of pregnant patients and contributes to an understanding of how to best manage and intervene on IE in pregnancy.

## Data Availability

Data was obtained through chart review and did not use any public databases.

## Acknowledgments

This area is to acknowledge contributions to the manuscript. All persons acknowledged must have seen and approved mention of their names in the article, as readers may infer their endorsement of data and conclusions; as such, acknowledged individuals must provide written (email) acknowledgment of their agreement to be included before acceptance. Emails from persons acknowledged can be forwarded to the editorial office or sent directly from the person. Please ensure the manuscript number is included in all correspondence.

## Sources of Funding

None

## Disclosures

The authors have nothing to disclose.

## Notes

### Competing Interest Statement

The authors have declared no competing interest.

### Author Declarations

This study was approved via the University of Pittsburgh Medical Center Internal Review Board (STUDY21060177)

